# Mobile Monitoring of Mood (MoMo-Mood) Pilot: A Longitudinal, Multi-Sensor Digital Phenotyping Study of Patients with Major Depressive Disorder and Healthy Controls

**DOI:** 10.1101/2020.11.02.20222919

**Authors:** Ana María Triana, Annasofia Martikkala, Ilya Baryshnikov, Roope Heikkilä, Tuomas Alakörkkö, Richard K. Darst, Jesper Ekelund, Erkki Isometsä, Talayeh Aledavood

## Abstract

Mental disorders are a major global cause of morbidity and mortality. The surge in adoption of smartphones and other wearable devices has made it possible to use the data generated by them for clinical purposes. In particular, in psychiatry, detailed and high-resolution information on patient’s state, mood, and behavior can significantly improve the assessment, diagnosis and the treatment of patients. However, there is long path to turn the raw data created by these sensors, to information and insights that can be applied in clinical practice.

Here, we introduce the MoMo-Mood Pilot: a study created to investigate the feasibility of using smartphones and wearables as data collection tools from subjects suffering from major depressive disorder. We collect data from 14 patients and 22 controls in two phases (active and passive). We demonstrate the feasibility of monitoring patients with several devices over short periods and passively monitoring them over long periods of time with minimal disruption in their daily activities. We identify and describe a series of challenges in this process.

The MoMo-Mood pilot study is an encouraging step in the process of determining the effectiveness of using wearables for quantifying the behavior and the state of psychiatric patients with high temporal resolution, which can lead to their potential adoption in clinical practice.

## 1 Introduction

Mood disorders, such as depression and bipolar disorder, are one of the main causes of the overall disease burden worldwide [Kassebaum et al., 2016]. Globally, depression affects 264 million people, while bipolar disorder impacts 45 million [James et al., 2018]. Psychiatric assessments commonly rely on interviews with patients and their relatives and suffer from memory biases. In addition to that, mental disorders lack clear biomarkers which can be measured in the clinic.

New technologies such as smartphones and fitness trackers create high resolution data on their users’ behaviors and physiology. Harnessing these data, and extracting patterns which are meaningful for clinicians, can help to improve psychiatric assessments and to find new behavioral markers for mental disorders.

Using consumer devices to passively collect objective data about patients’ routine could facilitate continuous assessment of behavioral manifestations relevant to psychiatric disorders. Such manifestations could be detected via signals like location, word sentiment, and social activity [Reinertsen and Clifford, 2018].

Here we introduce the Mobile Monitoring of Mood (MoMo-Mood) Pilot study, which aims to investigate the validity, feasibility, and utility of mobile phone applications for extracting clinically meaningful behavioral markers which can ultimately be incorporated and used for diagnosis and monitoring of mood state of patients with major depressive disorder (MDD). We analyze the data collected passively from three devices: smartphones, actigraphs, and bed sensors, and group the data into categories which are relevant to clinical symptoms. These categories are: sociability and phone activity, mobility, and sleep. We show that using smartphones and wearables for studying mental disorders is feasible and clinically relevant behavioral information can be extracted from the collected data. We also identify several challenges and bottlenecks that we encountered and reflect on our experience. These results and lessons are important for guiding future researchers in similar or larger-scale studies.

## 2 Methods

### 2.1 Recruitment of Study Participants

In this study two groups of participants, patients with Major Depressive Disorder (MDD) and healthy controls (HC), were recruited in Helsinki region, Finland. The recruitment was on a rolling basis and started in February 2017. The HC cohort were predominantly recruited through students’ mailing lists, while patients were all under treatment as outpatients in a university affiliated psychiatric clinic. During the initial meeting, the eligibility of the participants was confirmed by the researcher by using the Patient Health Questionnaire (PHQ-9). If eligible, the participant was given the detailed study information and consented. They would then fill out a series of detailed questionnaires (described in section 2.3.1). A data collection application would be installed on their phone and linked to the study server. They would also receive two additional devices on loan: a bed sensor and an actigraphy device. This marked the beginning of the first phase of the study (active phase) which lasted for two weeks. In this phase, active participation (answering to daily questions on the phone and using the loaned devices) was required. After this phase the participant met with the researcher again, returned the devices and the passive phase of the study which lasted 6 months for patients or one year (for controls) started. Patients however, could also continue to submit data for up to a year. During the passive phase participants were asked to keep the app installed, and answer monthly questionnaires (PHQ-9) on their phone, but otherwise have their usual activities. The participants were free to resign at any point in time. Details about the devices, the data collection app on the phone, questionnaires, and the study platform are described in subsequent sections.

### 2.2 Study Participants

For this study a total of 23 healthy controls and 14 patients with MDD were recruited. One control was not enrolled in the study for having a PHQ-9 score which fell in the sub-clinical depression range. The healthy controls were 17 females and 5 males, with the average age of 30.5 years (std: 10.4). The patients were 8 females and 5 males. The average age for the patients was 32.4 years (std: 12.2). One patient had not entered their age and sex information in the forms. 9 of the healthy controls were students, 7 were working, and 6 were working and studying simultaneously. 3 of the patients were studying, 3 were employed, and 8 were either not employed or on sick leave.

### 2.3 Data Sources

#### 2.3.1 Questionnaires

In clinical practice, the state of the patient is assessed by means of different questionnaires and interviews. We use well-established questionnaires to have a ground truth measurement of different variables (such as sleep, sociability, mood) which we can later compare with data that we gather from various sensors. We also use questionnaires to have a thorough evaluation of the patient and their psychological attributes. The baseline questionnaires that we used are: PHQ-9, Perceived Social Support Scale-Revised (PSSS-R), Morning Evening Questionnaire (MEQ), Adult ADHD Self-Report Scale (ASRS), Overall Anxiety Severity and Impairment Scale (OASIS) and NEO Five Factor Inventory (NEO-FFI).

#### 2.3.2 Smartphone

We used the AWARE phone application [Ferreira et al., 2015] to collect data from the phones. We collected communication (call and text) timestamps as well as an anonymous identifier for who the person has been in contact with, timestamp of phone screen on, off, lock and unlock events, GPS data, application use, levels of ambient noise, and battery status. We also used the phone to ask questions from the subject several times a day.

#### 2.3.3 ESM

*ESM* (Experience Sampling) is a method that has been long used in clinical studies to measure the emotional state of the person in-real time [Shiffman et al., 2008]. In this method usually the participant is prompted to answer questions about their state on their phone multiple times a day. For this study we used a shorter version of an *ESM* questionnaire which was developed for a previous study [Komulainen et al., 2014]. The questions showed up 3 times during the day (at random times within a certain time range) on the participants’ phones. In addition to that they received a series of questions in the morning to assess the previous night’s sleep, and another set in the evening to assess how the day was spent.

#### 2.3.4 Actigraph

Actigraphs are wrist-worn devices which are commonly used in clinical studies (including psychiatric studies) to measure participants’ activity levels and sleep [Tazawa et al., 2019]. In this study we used Philips Actiwatch 2 devices which could be used (without need for recharging) for two weeks.

#### 2.3.5 Bed sensor

For this study we used ballistocardiography-based Murata SCA11H nodes (bed sensors), which measure the amount of acceleration of the beds they are placed at. The participants were given the bed sensor together with a WiFi router which was pre-configured so that the bed sensor would connect to it and the collected data would directly be transmitted to the study server. The bed sensors locally analyze the data and as output provide heart rate, heart rate variability, respiration rate, stroke volume and signal strength at 1 Hz frequency.

### 2.4 Data Collection Platform

The Non-Intrusive Individual Monitoring Architecture (Niima) is a data collection platform which is designed and developed to be used for MoMo-Mood Pilot and other digital phenotyping studies [Aledavood et al., 2017]. This platform makes it possible to design studies with different parameters and link different types of data from various sensors. It also provides basic cleaning and pre-processing of the data to turn all data in standard tabular formats to make them easy to handle and analyze.

### 2.5 Privacy and Ethics of Data Collection

Our study was approved by the ethics committee of the Helsinki and Uusimaa Hospital District (HUS), and was granted research permit by HUS Psychiatry. The approval included data streams, platform data security, and information presented to subjects for consent. No data passed through any third-party platforms in the course of this study. All data in transit was encrypted. Written data security statement was approved both by local IT support and the research ethics committee. Also, participant privacy - even from study administrators - was a design value from the very beginning.

## 3 Findings

### 3.1 Dataset Description

Study participants provided different amounts of data and for different lengths of time (Table 1). Out of 46 subjects, 35 provided sleep sensor data, 33 actigraph data, 27 application, GPS, screen, and battery data, and 26 provided data from *ESM*, call logs, SMS, and ambient noise. The main reason behind the incompleteness of data sources was incompatibility of the data collection application with some of the subjects’ smartphones. All subjects were able to use the actigraph without problems, however we faced issues with the configuration of the devices at one stage which led to 3 subjects not receiving them at the time of enrollment. Out of all subjects, only one patient was unable to provide bed sensor data, because they did not have a stable place to live at the time of the study.

**Table 1:** Data summary per number of days. We show, for each data source, the mean and standard deviation of the overall number of days per group. Above the dashed line, we show sensors that collected data only during the active phase.

| data source | controls |  | patients |  |
| --- | --- | --- | --- | --- |
|  | Mean | SD | Mean | SD |
| ESM | 16.61 | 5.11 | 14.75 | 4.29 |
| actigraph | 13.89 | 0.31 | 13.21 | 1.42 |
| sleep sensor | 15.36 | 5.33 | 16.31 | 8.15 |
| application | 123.28 | 100.85 | 89.44 | 114.63 |
| screen | 134.94 | 100.66 | 102.67 | 115.94 |
| battery | 135.28 | 99.50 | 103.11 | 115.97 |
| call logs | 84.22 | 62.78 | 69.00 | 73.91 |
| SMS | 66.22 | 57.82 | 61.13 | 66.67 |
| GPS | 107.72 | 93.09 | 89.00 | 96.57 |
| ambient noise | 115.06 | 89.74 | 115.36 | 116.80 |

Patients had an average PHQ-9 score of 13.9 (std: 6.19) and average OASIS score of 12.86 (std: 3.30). PHQ-9 scores higher than 10 means that diagnosis of depression is likely. In our pilot, most patients had mild depression at the time of assessments, 4 patients had moderate depression, and 3 people had severe depression. OASIS scores higher than 6 means likelihood of at least some impairment due to anxiety, while scores higher than 8 are indicative of anxiety disorders. So, the included patients were relatively more anxious than depressed. Thus, the included sample has a lot of variation in terms of the severity of symptoms.

### 3.2 Sociability and Phone Activity

We begin the descriptive analysis of the data by comparing the mean subject’s daily averages per group for each sensor; i.e. for each subject, we calculate the daily average, and then take the mean over the days. One patient was discarded because they only have 4 seconds of data. 3 controls were discarded because their data needed special pre-processing (different operating system).

We observe that controls and patients unlock their phones a similar number of times (Fig. 1 A). However, the median patient’s screen is on in less occasions than the median control’s screen. This suggests that in our sample the median patient receives less notifications or presses the power button less than the median control. Further, the median patient’s screen is on and unlocked for longer periods of time than the median control’s screen (Fig. 1 B). This suggests that our median patient uses the phone for longer than the median control, despite unlocking the phone a similar number of times.

**Figure 1:**
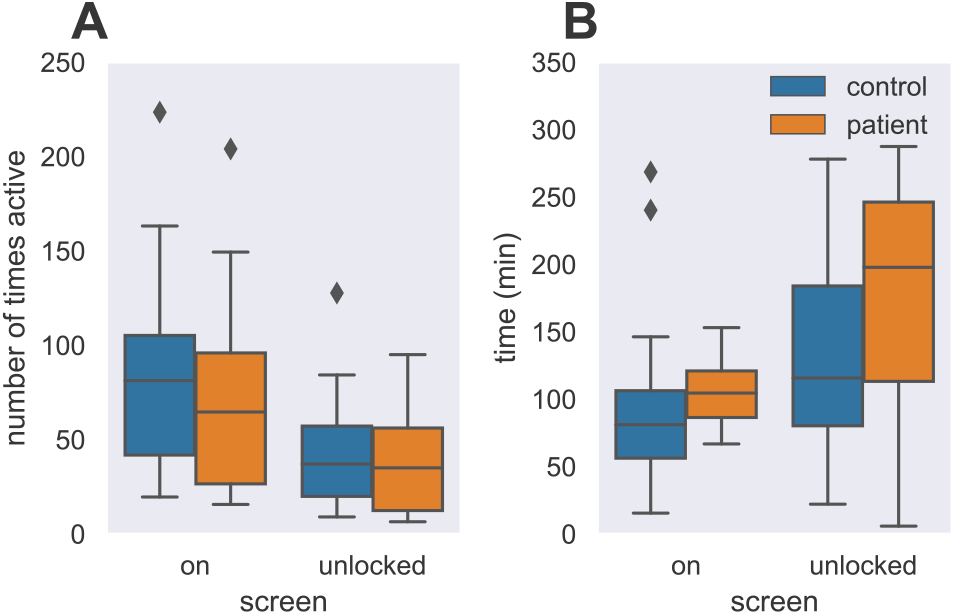
Screen use patterns per group. Controls and patients use their devices with different patterns. (A) Although controls and patients unlock their phones a similar number of times, the median patient’s phone screen is on in less occasions than the median control’s phone screen. (B) The median patient unlocks the screen for a longer time than the median control. Similarly, the median patient’s screen is on for a longer time than the median control’s screen.

Next, we examined what kind of apps the subjects used. Apps were manually classified into 9 different categories according to their main purpose, following the Android Play Store classification. 3 app categories were rarely used: transportation, travel, and productivity. Figure 2 shows the duration per group of the 7 most frequent app categories. We see that on average, subjects spend less than 25 minutes per day in sport, shops, and games apps with two exceptions (Fig. 2 A). These special cases are one control who used a step counter app, and another control who played a strategy game approximately 3 hours per day. Few subjects read/watch the news on their phones, but 2 of them spend more than 1 hour on a daily average (see Fig. 2 B). Moreover, we note that the median patient employs slightly more time using entertainment and social media apps than the median control (Fig. 2 B), although they spend a similar time in communication apps (see Fig. 2 C). For all cases, the patient group has a larger spread than the control group.

**Figure 2:**
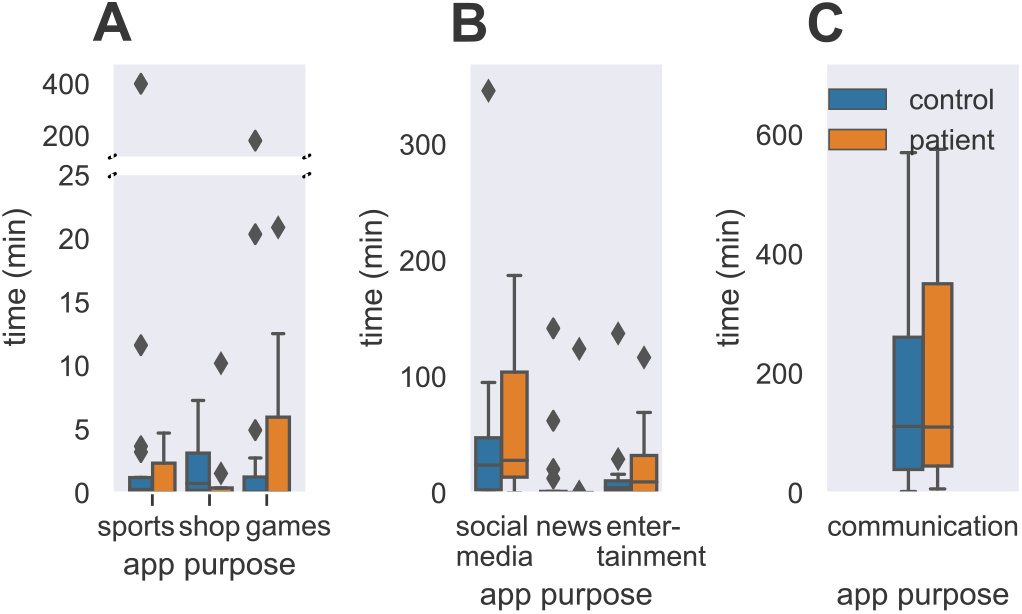
Mobile apps usage per group. Subjects use the mobile apps differently according to their purpose and groups. (A) Subjects rarely used mobile apps for sports, shopping, and games, except for two controls who used the phone for step counting and strategy games. (B) Few subjects used their phones to read the news and for entertainment, but more time was allocated to watch social media content. The median control uses social media apps for a similar duration than the median patient, but patients have a higher variation within their group compared to the controls. (C) The median patient spends a similar amount of time in communication apps than the median control, despite this time varies higher within patients than within controls.

Finally, Fig. 3 shows the calls and SMS statistics per groups. Here, we see that the median patient has more incoming and outgoing SMS than the median control (Fig. 3 A). The median patient also receives and misses more calls than the median control, but places a similar number of calls (Fig. 3 B). Similarly, the median patient’s calls are longer than the median control’s, regardless of the category or contacts (Fig. 3 C). This is an interesting pattern, although we can only wonder if the higher number, longer incoming calls are due to friends, relatives or health care providers to check up on the patients. Unsurprisingly, subjects devote more time to new means of communication, like apps and social media, than traditional methods like calls (Fig. 3 D).

**Figure 3:**
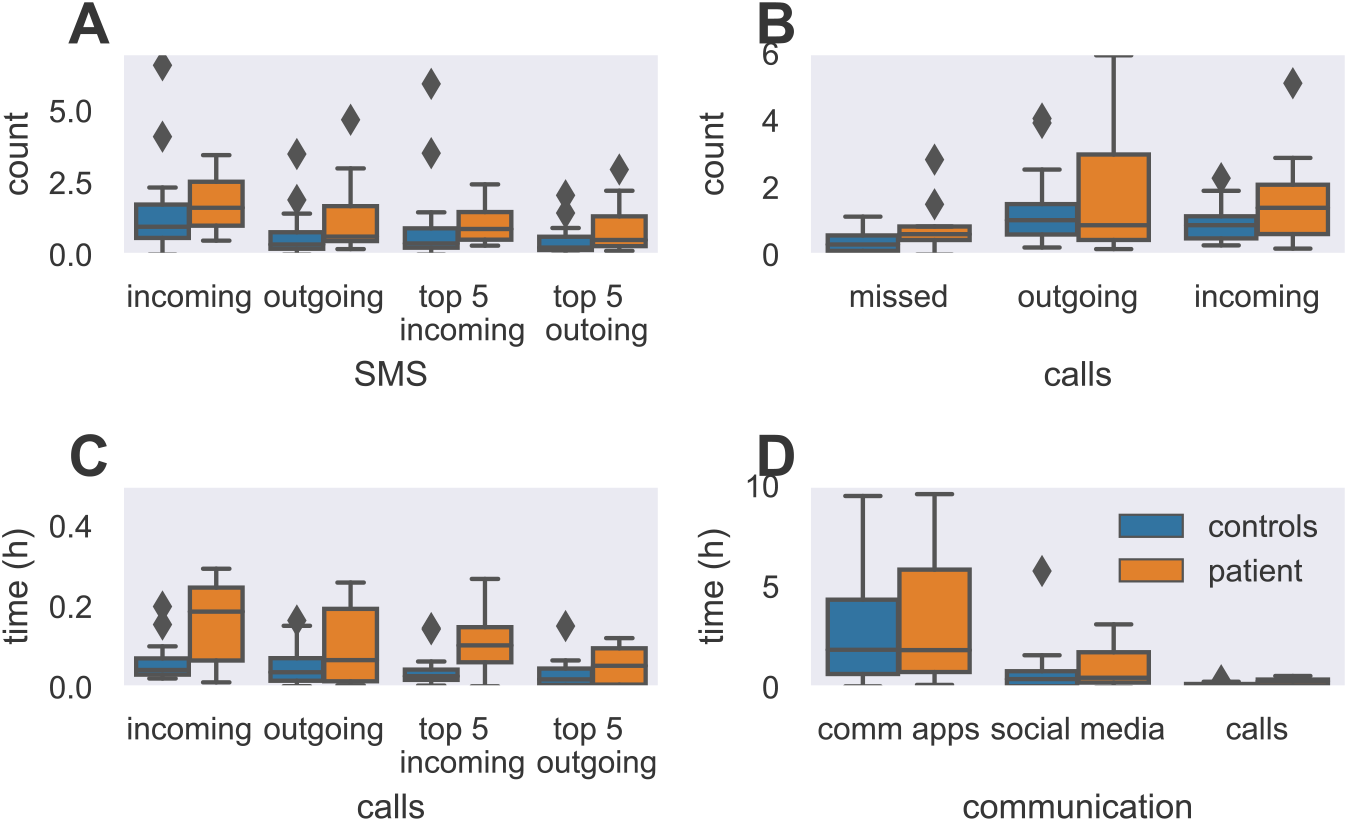
Communication patterns per group. Subjects use the mobile devices for communication purposes differently according to their group. We see that: (A) the median patient uses more SMS than the median control. However, the number of received and sent SMS varies more for patients than for controls. We also see these patterns in messages from/to the 5-most frequent contacts (top 5 incoming and top 5 outgoing). (B) The median patient places and misses a higher number of calls than the median control. Controls have less variation in their number of incoming and outgoing calls than patients. (C) The median patient spends more time in their calls than the median control. However, this time has a higher variation in patients than in controls, as seen in the wider spread of the boxplots. This is also visible when we only analyze the incoming and outgoing calls of the 5-most frequent contacts. (D) Controls and patients spend similar median time on their phones using communication apps. However, compared to controls, patients have a higher variation in the number of hours spent on these platforms and on social media. We also notice that most of the communication is done by using apps rather than calls.

### 3.3 Mobility

Next, we employ a descriptive analysis on the movement data from GPS and actigraph (Fig. 4). We notice that in our sample, the median patient moves fewer hours than the median control (Fig. 4 B). This is supported by previous literature in which low energy is associated with depressive symptoms [Arnold, 2008]. However, we do not observe the same pattern from the GPS data. On the contrary, GPS data suggest that the median control and median patient move for almost the same time. Nevertheless, it is important to remember that these two sources measure different aspects of movement and while a subject wears the actigraph continuously, they may not carry their smartphone with them at all times.

**Figure 4:**
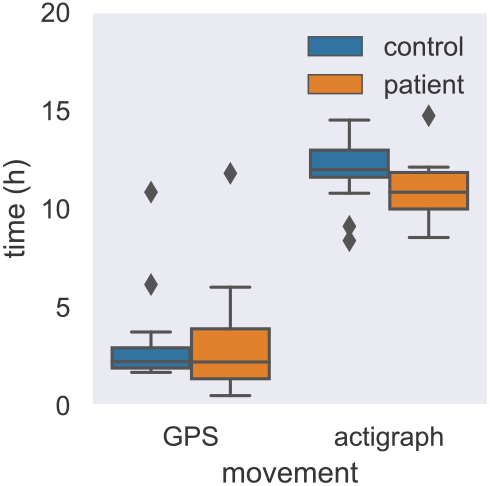
Mobility patterns per group. The median patient spent less time moving than the median control, as shown by actigraph. However, this pattern is not visible for the median subject in the GPS data.

### 3.4 Sleep

Finally, we compare data from the actigraph and sleep sensor. We discard one subject because they had only 3 days of low-quality data. We observe that the median patient spends longer hours resting than the median control, as measured by both devices (Fig. 5 A). We can also notice a difference in the time spent measured by actigraph and bed sensor for all groups. This can be related to differences in the rest-detection algorithms of the devices. Fig. 5 B shows the fraction of sleep out of the total rest duration. We notice this ratio is closer to 1 in controls, i.e. controls are mostly sleeping if they are resting, but patients might have longer resting time without sleeping.

**Figure 5:**
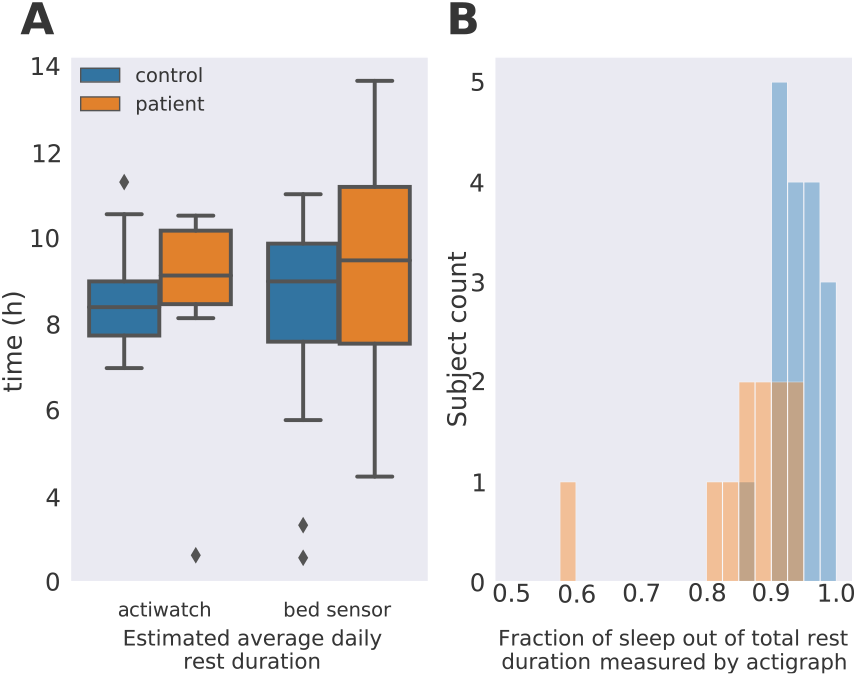
Resting patterns per group. We computed the average resting times based on actigraph and bed sensor data. (A) In this dataset, the median patient spends longer hours resting than the median control. (B) Actigraph rest and sleep data for patients and controls.

## 4 Challenges and Insights

### 4.1 Technical Challenges

We faced 3 technical challenges: (1) how to reliably collect data from smartphones in a non-intrusive way, (2) how to organize, store, and manage the longitudinal data coming from different subjects and sensors, and (3) how personal phone settings may impact the data.

Currently, there are several mobile apps that collect data from smartphone sensors. However, we found problems with their stability and support. For example, some apps crashed after 15 days of continuous data collection and we could not establish contact with the developers to learn why. After careful consideration, we decided to deploy the AWARE framework app which performed best in our tests. Still, 5 controls and 2 patients reported it malfunctioned on their phones.

To overcome the second challenge, we developed Niima (see section 2.4) and linked the AWARE app. Although AWARE has its own data collection platform, it did not allow the integration of the other devices included in the pilot. Initially, we could only collect data via the AWARE Android app. The AWARE iOS app was incorporated later on (3 controls). However, we could not collect *ESM* data (see section 2.3.3).

During data exploration, we learned that information provided by phones vary with brand and operating system (OS). This affects the generalization of the pre-processing strategies and the analysis of missing data points. Moreover, users have different settings that affect how much the app can track. Thus, even for one subject, data from distinct smartphone sensors may vary. For example we observe one subject’s settings prevented the app to track ambient noise.

In general, technical challenges are laborious and may demand some additional resources, even after the data collection phase.

### 4.2 Enrollment and Adherence Challenges

Due to the nature of this study, the initial screening for the enrollment cannot be fully self-administered. This design limits the number of subjects that can be recruited per day. Patients, were always screened and enrolled by a psychiatrist which made the enrollment heavily dependent on the available time by the psychiatrist. On average, patients also had a higher tendency to cancel or move the time of the meeting with a short notice.

On average, we collected 17.25 days per subject during the active phase. Similarly, during that phase, all subjects collected data from one or more sources and 23 people have a 100% compliance score across all streams. In our pilot, 68% of the controls and 43% of the patients decided to continue with the passive phase. Half of these subjects stopped data collection before 110 days and 6 subjects have gathered more than 250 days.

Keeping subjects motivated during both phases is a major challenge. We provided our healthy controls (patients) with 4 (2) movie tickets. However, they were given all tickets by the end of the active phase and did not have any additional compensation. Future studies can provide motivators – like visualization of subject’s data – to improve adherence in the passive phase. Also, systems can be implemented so that the subject is contacted in case of sudden disruption of incoming data.

External factors can also threat the continuity of data collection. For example, Android has restricted the use of many permissions that the current AWARE app employs; therefore,it cannot be downloaded from the play store, which is the subjects’ most familiar method to install apps.

### 4.3 Privacy and Data Handling Challenges

Subject’s privacy was a key concern in this study due to the sensitive nature of the collected data. We face this challenge in two levels, technology and human.

On the technology part, all data is collected by the Niima platform and linked by a randomly generated subject identifier. Raw data is processed for privacy and re-assigned new identifiers. Only then, data can be extracted for follow-up analysis on a centralized computing cluster. For example, no GPS coordinates are available to researchers; instead researchers have access to aggregated, derived data like total distance per day. No data passed through any third-party platforms and all data in transit was encrypted. Only actigraph data had to be manually uploaded to Niima.

On the human side, two teams of researchers worked in the pilot. The first team conducted the interviews and met the subjects, while the second one analyzed the data streams. This means that each team only had access to part of the information, but not all. The researchers meeting subjects cannot extract any data from the platform and the researchers analyzing the data cannot know who the subjects are. Inevitably, some highly sensible information was collected for a brief period (e.g. contact information). These data were handled using the same protocols employed in medical care.

### 4.4 Data Completeness

For this pilot, we have two different types of sensors, depending on their sampling frequency (fixed or variable). Fixed sensors are sources that are continuous or have a fixed sampling rate that we know. Variable sensors use event-sampling approaches. Thus, we can easily detect missing data points for fixed, but not for variable sensors; i.e. we need to create strategies that can detect missing data for variable sensors. At this point, combining distinct sources of information is crucial as they provide different details of the same period. By merging the two sensors events, we are guaranteeing to have a more complete scenario.

We have learnt that depending on the research question and data source, we can tolerate higher levels of missing data. Still the question remains, what is a good amount of data per subject? Although there are methods to solve this problem, we still need to think about how we perceive missing data. Is it some undesirable consequence or does it have meaning? In this case, missing data might indicate behaviors of fatigue and social withdrawal which are symptoms related to MDD. Here, the most important lesson is that we need to keep a flexible mindset to understand and handle missing data.

### 4.5 Challenges Related to Human Behavior

Monitoring people in real life scenarios is demanding. Every person has different life circumstances that may jeopardize the quality and continuity data collection. These events can cause data gaps in the data or interruption in incoming data. Although normal, data-gaps events may make the data interpretation more difficult. Handling missing data from patients is additionally delicate, because data gaps (e.g. communication events) could be telling of worsening of the patient’s state. Therefore, further investigation to identify reasons behind longer data gaps are necessary. We need to find out whether these gaps are related to technical issues, external life circumstances of the person, or changes in the internal states and mood. Complimentary questions about daily and life events may help solve this problem. In addition to that, combining data from different devices could provide additional insights about the underlying reasons.

## 5 Discussion

In this pilot study, we investigated the feasibility of using smartphones and wearables as data collection tools from subjects suffering from major depressive disorder. We gathered data from 22 controls and 14 patients. Data is collected from multiple devices (smartphones, actigraphs, and bed sensors) across different periods of time.

Our results show that: (1) longitudinal, multi-source data collection projects are feasible including controls and patients populations, (2) subjects interact differently with their smartphone, and (3) patients’ measured behavior is more spread, suggesting the heterogeneity of the disorder symptoms can be captured by mobiles and wearables. However, due its pilot nature, we cannot generalize the preliminary results since there could be some underlying, simpler explanations for the observed behaviors. For example, the wide spreads of the groups distributions that may prevent any significant statistical inferences in this sample.

We also identified 5 areas of challenges that future researchers should take into account. Such areas are challenges related to technical, enrollment and adherence, privacy and data handling, data completeness, and human behavior. We learned 4 lessons about these type of studies: (1) they demand considerable efforts from multidisciplinary sources and therefore, keeping organized records of information in an unified system is crucial; (2) they require substantial efforts in technical development given the multiple sources of data; (3) they need holistic approaches for analyzing the data because distinct sources provide complementary information about an event; and (4) they require researchers to be flexible and adapt to unforeseen situations as the data collection progresses.

Although this is a rather small (in terms of the number of participants) feasibility study, we observe some interesting group patterns. It seems like depressive subjects are less physically active than controls and also spend more time resting. Interestingly they spend more time in phone calls, indicating a greater interest in inter-subject communication which is rather surprising. We do not have data on the subject’s time spent physically meeting others. Finally the depressed subjects spend more time using social media related applications than healthy controls. Such findings need to be examined in larger sample sizes, which is a clear limitation in our pilot. In our case, the rather small sample size prevents any significant result with respect to group differences to be derived. Understanding the causality of the group-wise difference in social media usage would require a prospective study design.

These findings are encouraging as they represent the preliminary data exploration that will help building a larger, more comprehensive study. We hope this pilot serves as a guideline for future projects that contribute to the advance of understanding and treating mental health problems.

## Data Availability

The data is not openly available.

## 6 Acknowledgements

AT and TA acknowledge support from the Academy of Finland, project “Digital Daily Rhythms", No. 297195. TA acknowledges support from the James S. McDonnell Foundation. We also acknowledge the computational resources provided by the Aalto Science-IT project.

